# The effect of population migration on the diffusion of cholera outbreaks in metapopulations

**DOI:** 10.1101/2022.04.12.22273810

**Authors:** Mahir Demir

## Abstract

In this study, an improved Susceptible-Infected-Recovered (SIR) epidemic diffusion model for cholera is extended by including migration for susceptible people. This model is applied to a metapopulation that consists of two isolated cities where just susceptible individuals can migrate between the cities. The disease-free equilibrium, the endemic equilibrium points, and the basic reproductive number with unequal migration rates are analyzed for this metapopulation. Firstly, the study showed that the basic reproductive number depends on the migration rates between the cities. Then, showed that when the epidemic SIR system is stable, then the infected cases for cholera outbreak can reach zero in one city, but the infected cases in the other city still can stay positive. Finally, discussed three scenarios that depend on population sizes and migration rates of susceptible people between the cities and showed how important the migration rates are in the diffusion of the cholera outbreak by visualizing these three scenarios.

**Mathematics Subject Classification:** Primary: 92B05; Secondary: 92D40

## 1 Introduction and Background

Cholera is a bacterial disease usually spread through contaminated food and water, and it causes severe diarrhea and dehydration. With no treatment, cholera can be fatal in a matter of hours, even in previously healthy people. The last major outbreak in Turkey occurred in 1970. But cholera is still present in Africa, Southeast Asia, Haiti, and central Mexico. The risk of a cholera epidemic is highest when poverty, war, or natural disasters force people to live in crowded conditions without adequate sanitation (Mayo Foundation for Medical Education and Research (2016)).

Studies of cholera outbreaks have been estimated that each year there are 1.3 to 4.0 million cases of cholera, and 21 000 to 143 000 deaths worldwide due to cholera, and most of those infected have no or mild symptoms and can be successfully treated with oral rehydration solution. Highlighted by the 2008 outbreak in Zimbabwe, a new focus on controlling cholera has emerged due to the rise in cholera incidence. The recent outbreaks have caused enormous loss of life and financial devastation to families and the healthcare system (World Health Organization (2008)).

Modeling on epidemic diffusion mostly concentrates on the compartment epidemic models of ordinary differential equations (Aslan et al. (2022); Demir et al. (2021)). In such a modeling framework, the total population is divided into several independent classes, and each class of individuals is closed into a compartment. To describe the mechanism of cholera disease, King et al. (2008) used a two-path cholera model including a class for severe infections as well as a class for mild infectious and showed that natural immunity to cholera may wane within a year. Samsuzzoha et al. (2010) used a diffusive epidemic model to describe the transmission of epidemics. The equations were solved numerically depending on the different initial population densities. Similarly, Shi and Dong (2012) formulated and discussed models for spreading infectious diseases with variable population sizes (Liu and Xian (2013)).

Besides the above studies, Miller Neilan et al. (2010) used a compartmental model and implemented an optimal control strategy in the control of cholera disease under different control variables. Such a control approach is applicable to many areas in ecology from fishery management to infectious diseases (Lenhart and Workman (2007); Demir and Lenhart, (2020); Demir and Lenhart, (2021)). Lee et al. (2012) extended an SEIR model to incorporate population migration between cities and investigated the effectiveness of travel restrictions as a control against the spread of influenza. In addition, Hethcore (1976) proposed deterministic communicable disease models by using a system of ordinary differential equations and considering population migration (only equal rates were considered), which led to different equilibrium points.

In this study, an improved Susceptible-Infected-Recovered (SIR) epidemic diffusion model for cholera (Miller Neilan et al. (2010)) is extended by including migration for susceptible people, and the effect of migration between two isolated cities is discussed to investigate the diffusion of cholera outbreaks. Firstly, the effect of population migration on the basic reproductive number obtained for two cities is discussed when a cholera outbreak occurs in these two cities. Then, the effect of migration between these cities on endemic equilibrium points in which the disease exists in one city and dies out in the other city is analyzed under different scenarios.

## 2 Epidemic Diffusion Model (SIR)

The objective of this study is to formulate a mathematical model for cholera, which migration can occur just between susceptible individuals in two isolated cities with unequal migration rates. Human populations are divided four different classes in both cities: Susceptible class(*S*_*i*_), Asymptomatic infected class 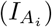, Symptomatic infected class 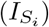, Recovered class (*R*_*i*_). Thus, the total number of individuals in city *i* is 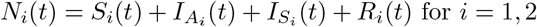., and the total number of individuals in both cities, *N* = *N*_1_ + *N*_2_. In the model just susceptible individuals can migrate with migration rate *a*_*i*_ (*a*_1_ *≠a*_2_). For each city, two classes of bacteria concentration are considered, one that is hyper-infectious class 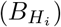 and one that is less-infectious class 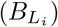. For the transitions among compartments and the description of parameter given for these transitions, see Figure 1 and Table 1.

**Table 1:** Descriptions of model parameters

| Parameter | Description |
| --- | --- |
| $p$ | Proportion of infectious being asymptomatic |
| $e_1$ | Death rate due to asymptomatic |
| $e_2$ | Death rate due to symptomatic |
| $\gamma_1$ | Recovery Rate of asymptomatic |
| $\gamma_2$ | Recovery Rate of symptomatic |
| $\eta_1$ | Shedding rate of asymptomatic |
| $\eta_2$ | Shedding rate of symptomatic |
| $\kappa_{H_i}$ | Half saturation constant of hyper-infectious |
| $\kappa_{L_i}$ | Half saturation constant of Less-infectious |
| $\beta_{H_i}$ | Ingestion rate of hyper-infectious |
| $\beta_{L_i}$ | Ingestion rate of less-infectious |
| $w$ | Immunity waning rate |
| $\delta$ | Bacteria death rate in $B_{L_i}$ |
| $\chi$ | Bacteria transition rate from $B_{H_i}$ to $B_{L_i}$ |
| $a_1$ | Migration rate from city 1 to city 2 |
| $a_2$ | Migration rate from city 2 to city 1 |

**Figure 1:**
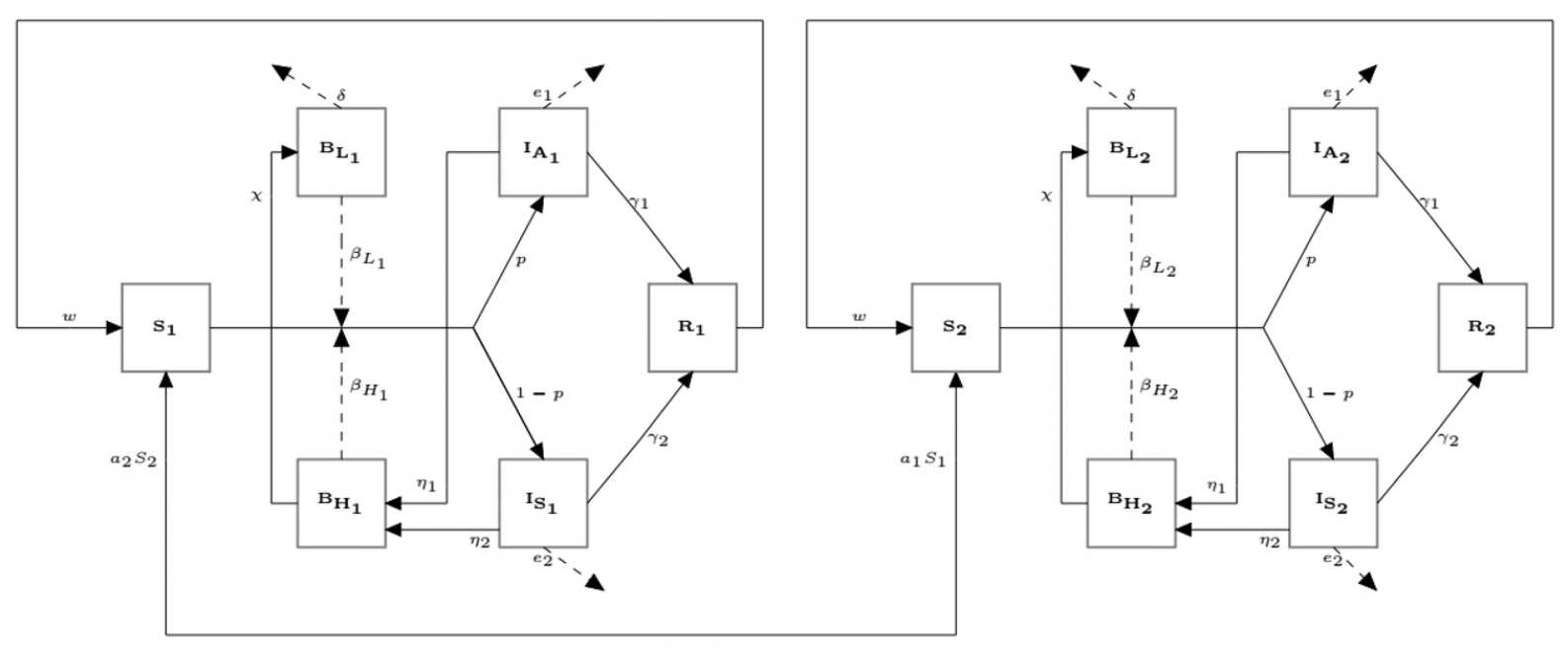
Flow diagram of model illustrating the disease transitions among the compartments (modified from Miller Neilan et al. (2010)).

### ODE system

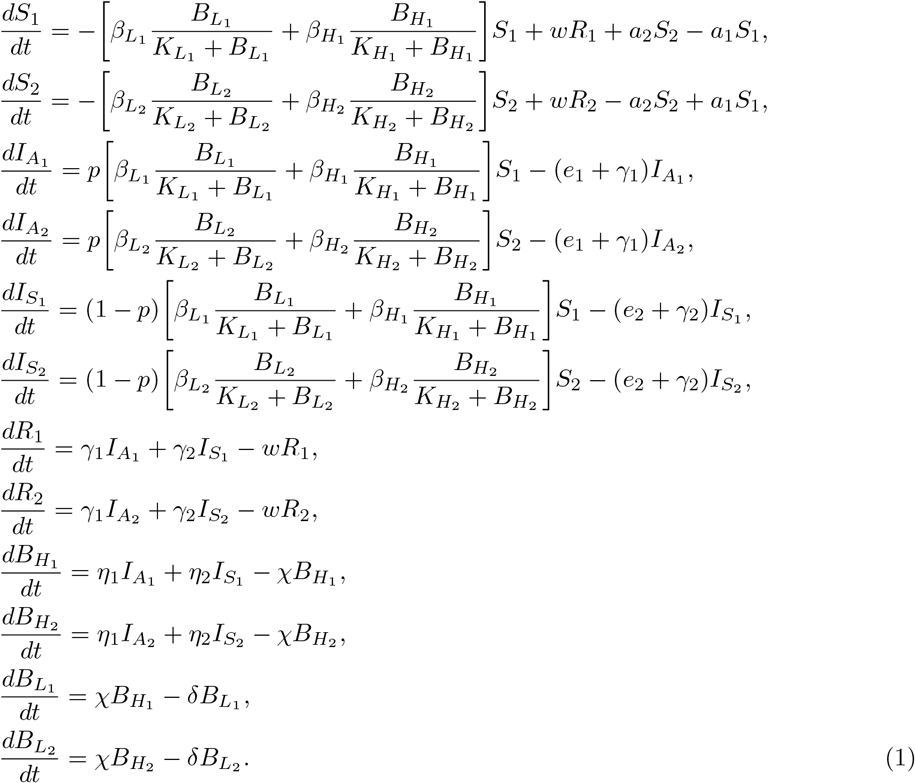

with the initial conditions: 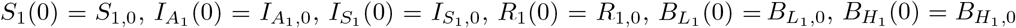, and 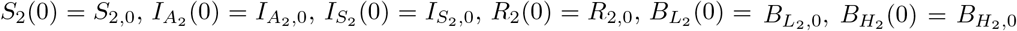. These all the initials and parameters given in the ODE system (1) are considered to be non-negative and bounded. The left-hand side of ODE system (1) represents the rate of change in the size of each compartment. Thus, in the ODE system (1), we have the following

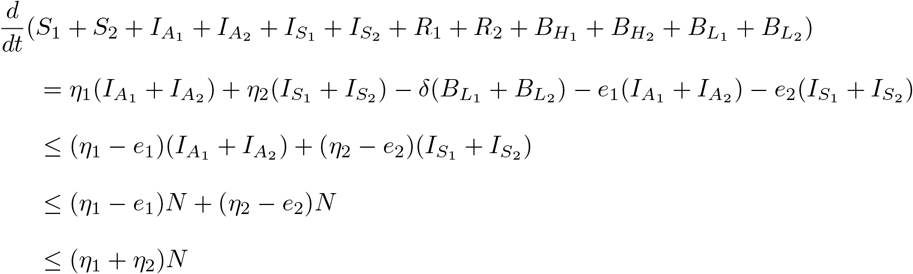

where *N* = *N*_1_ + *N*_2_ such that 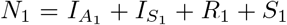 and 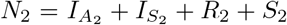. Thus, the ODE system (1) is bounded and so its solution is also bounded. Thus, the ODE system (1) is biologically feasible to be investigated with this modeling framework. When we substitute 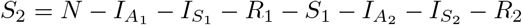 in the ODE system (1), we will get the following ODE system:

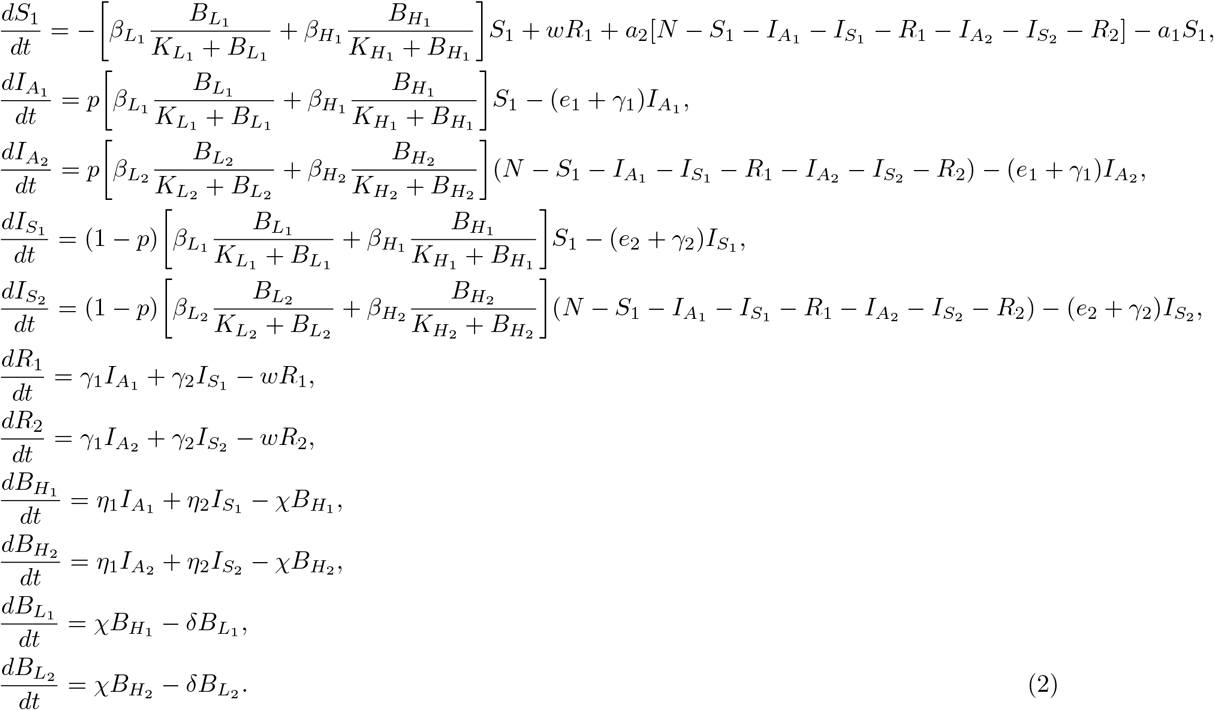

I reduce the number of equations for simplicity and I will consider the ODE system (2) in the rest of the analysis and obtain numerical results by using the ODE system (2).

## 3 The Basic Reproduction Number of the Metapopulation

To determine the basic reproductive number for the model given in (2), I will use the next generation matrix (Van den Driessche and Watmough, (2002)). Consider the system to be ordered such that 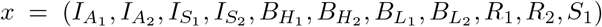. Here, our disease-free equilibrium point of the metapopulation is 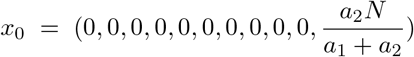 when we let 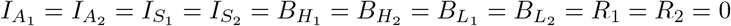.

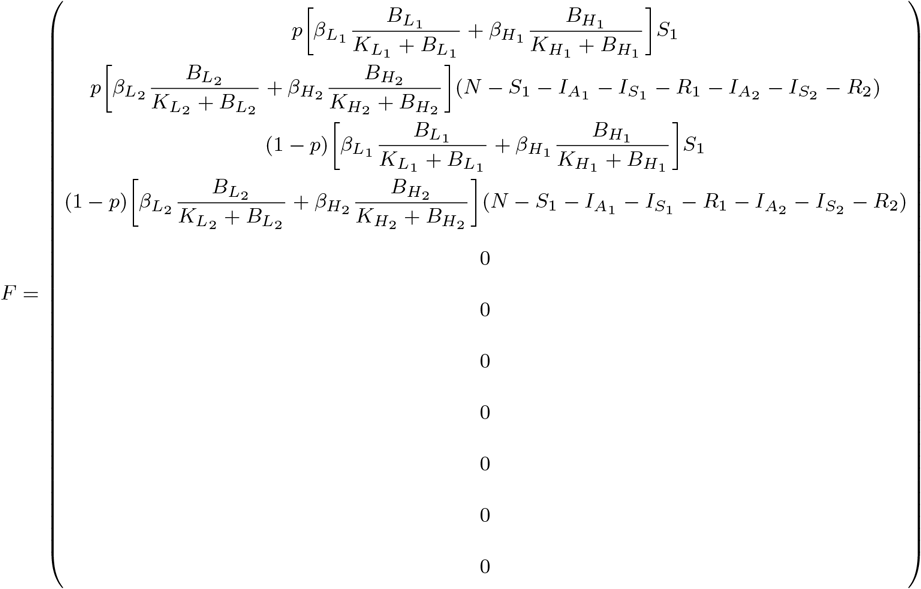

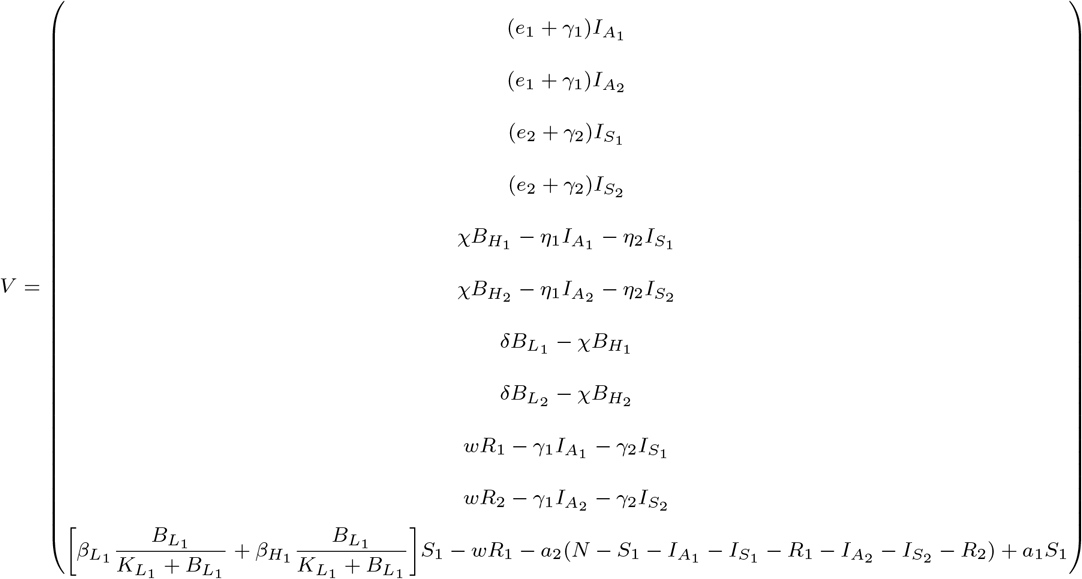

Compartments 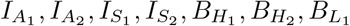, and 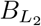 are considered to be the infectious compartments. Let’s define 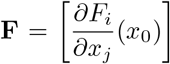 and 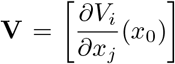 for 0 *≤ i, j ≤* 8. Here **F** is the Jacobi matrix of *F*, and **V** is the Jacobi matrix of *V* such that

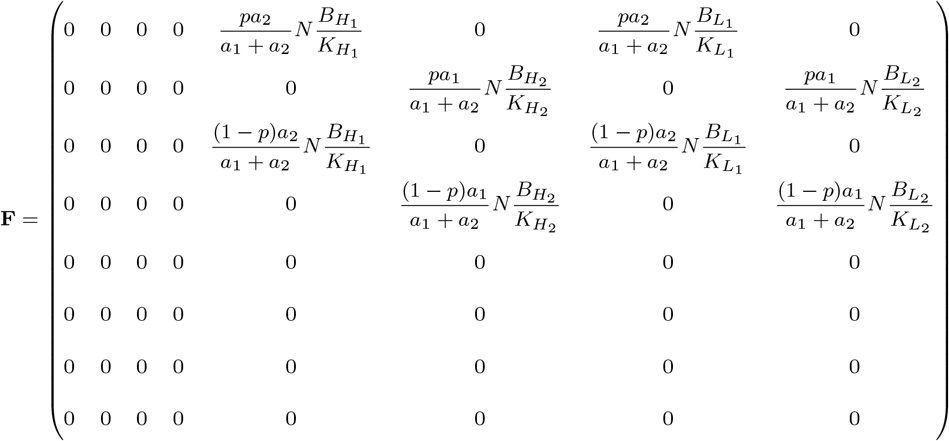

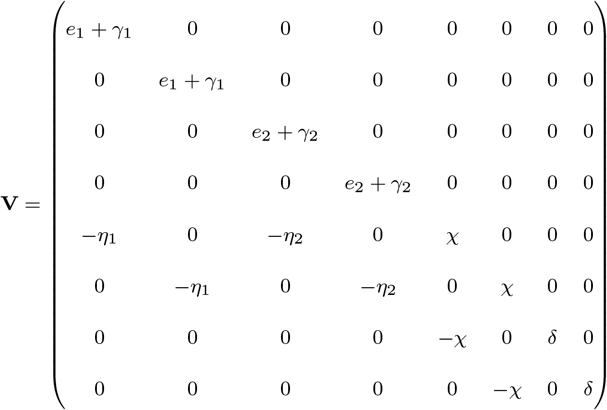

Now, we first calculate the *V* ^*−*1^ such that

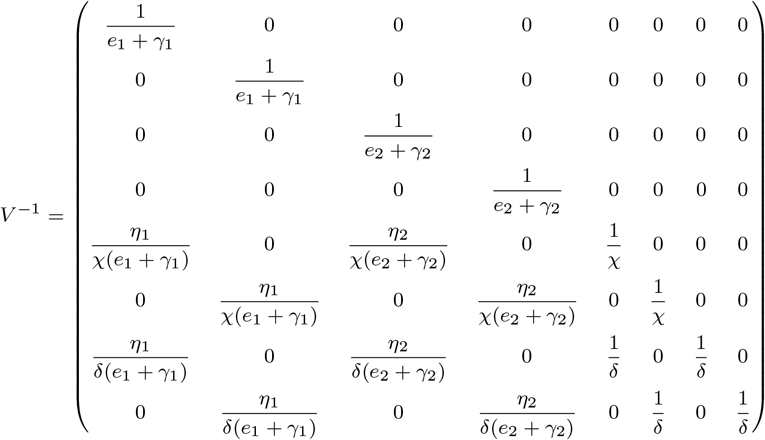

After re-scaling the parameters such that 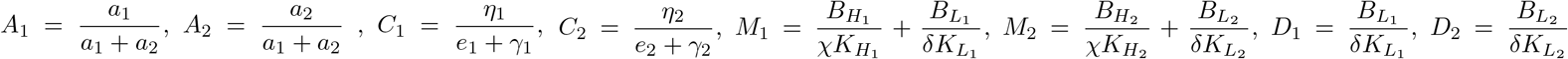 and *p*^*\**^ = 1 *− p*.

*FV* ^*−*1^ is obtained as

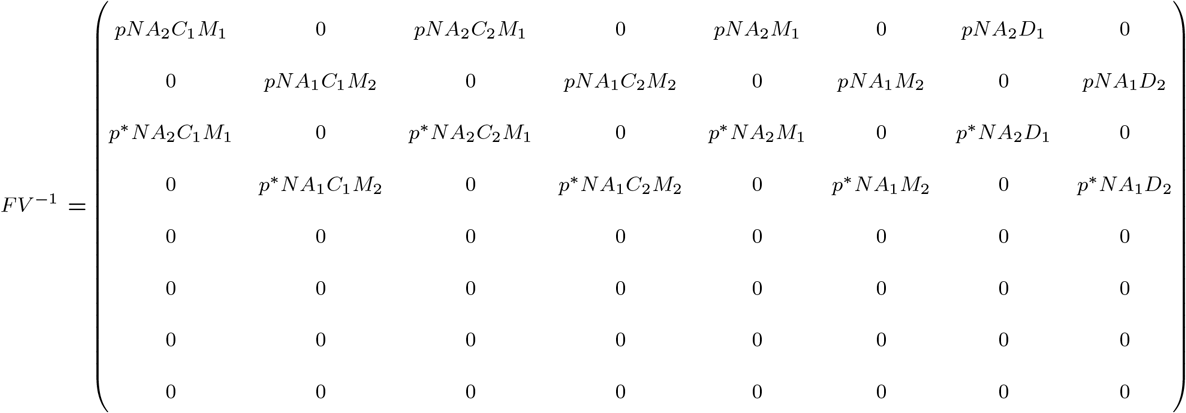

To be able to obtain the basic reproductive number *R*_0_ of the metapopulation, we need to find the dominant eigenvalue of the matrix, *FV* ^*−*1^ by setting it as *det*(*FV* ^*−*1^ *− λI*) = 0, where *λ* denotes eigenvalues and *I* denotes the unit matrix.

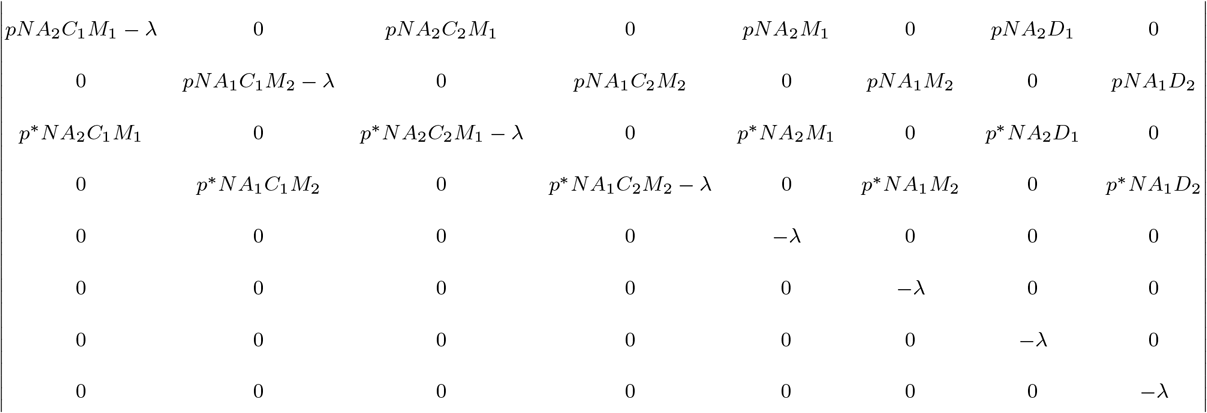

by solving this determinant, *R*_0_ is obtained as:

If *A*_2_*M*_1_ *≥ A*_1_*M*_2_ then

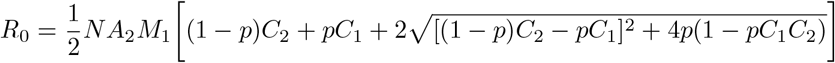

If *A*_1_*M*_2_ *≥ A*_2_*M*_1_ then

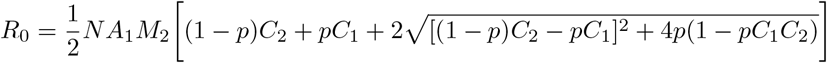

where 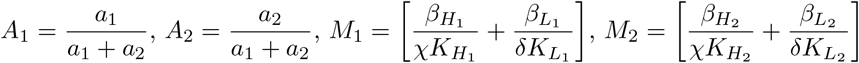.

Thus, after re-scaling back to the original parameters, we obtain the following

If 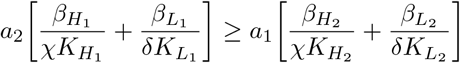 then

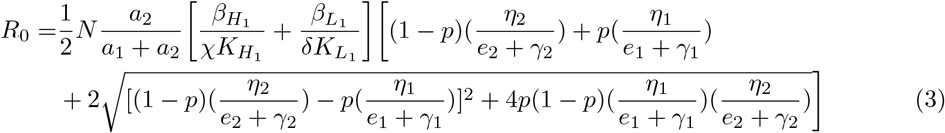

If 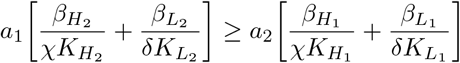 then

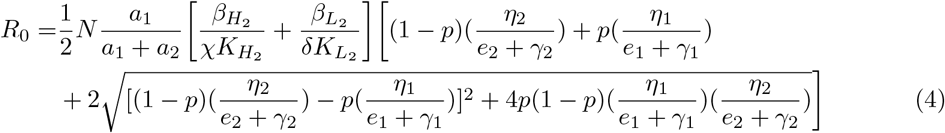

The basic reproductive number, *R*_0_ obtained for both cities depends on migration rates, *a*_1_ and *a*_2_ as well. Thus, the migration between these cities can not be ignored in the control of the cholera outbreak in these two cities. When the rate *a*_2_ increases in Eqn.3, then *R*_0_ increases. Similarly, any increases in the rate, *a*_1_ in Eqn.4 increases *R*_0_ as well.

## 4 Existence of the Equilibriums of the Metapopulation

### 4.1 The Disease-free Equilibrium Point of the Metapopulation

The disease free equilibrium point, where all the human population is susceptible is one of the key concepts in the investigation of outbreaks, especially for the calculation of the basic reproductive number *R*_0_. When we let 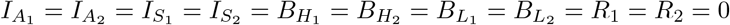, then we can get the equilibrium point 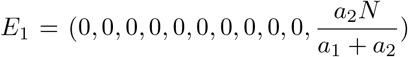. The number of infected individuals in both cities are zero, and these two cities just consist of susceptible individuals.

### 4.2 Endemic Equilibrium Points of the Metapopulation

In this part, an endemic equilibrium point means the cholera disease exists in city 1 and does not exist in city 2, or vice versa. When we let 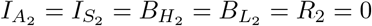 for city 2 and set the right hand side of the ODE system (2) to zero, then we can obtain the second equilibrium point which is an endemic equilibrium point, 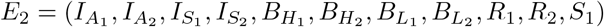 as below

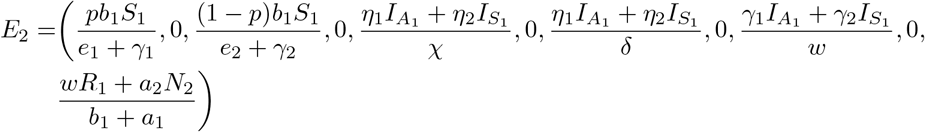

where 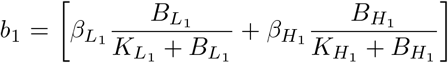. When the equilibrium point, *E*_2_, is stable, then the number of infected individuals in city 2 is zero, but for city 1 the number of infected individuals is still positive. Thus, we call this equilibrium point as an endemic equilibrium point. When we closely look at the equilibrium point *E*_2_, all the entries are positive in city 1 for 1 *> p*. Thus, this equilibrium point will stay positive for the city 1 until *S*_1_ class reaches to zero. When *S*_1_ class hits zero, almost all the people in city 1 will be in the recover class, *R*_1_. Note that the recover class, *R*_1_ depends on the migration rates *a*_1_ and *a*_2_ as well for this equilibrium point.

Now if we let 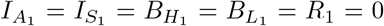 for city 1 and set the right hand side of the ODE system (2) to zero, then we can obtain the third equilibrium point of the Epidemic diffusion system, 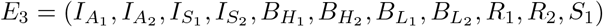 as below

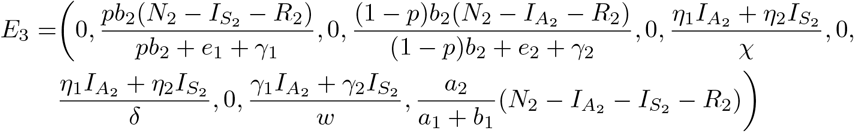

where 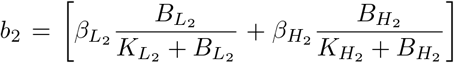. When we plug 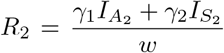 in *E*_3_, we will get the following

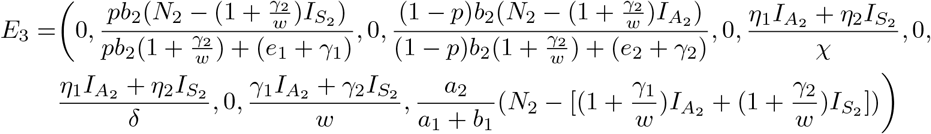

When the equilibrium point, *E*_3_, is stable, then this time the number of infected individuals is zero in city 1, but positive in city 2. For the conditions, 1 *> p* and 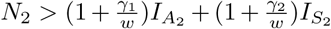, all the entries for city 2 will stay positive.

## 5 Stability Analysis of the Equilibrium Points

Now, I check the stability of equilibrium points. However, it is not easy to examine the stability analysis analytically for the ODE system (2). Therefore, I examine the stability analysis numerically but first, need to obtain the Jacobi matrix of the ODE system (2). Lets define 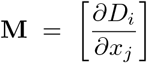 for 1 *≤ i, j ≤* 11. Here **M** is the Jacobi matrix of the ODE system (2). *D*_*i*_ for 1 *≤ i ≤* 11 are the right side of differential equations in the ODE system (2) and 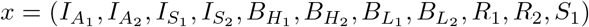. After obtaining the Jacobi matrix *M*, the equilibrium points *E*_1_, *E*_2_, and *E*_3_ are plugged into the Jacobi matrix *M* to check the stability of these equilibrium points. I conducted the stability analysis by using parameter values given in Table 2 and obtained that the disease-free equilibrium point is stable, but endemic equilibrium points are not.

**Table 2:** Descriptions and values of model parameters

| Parameter | Description | Value | Unit | Source |
| --- | --- | --- | --- | --- |
| $p$ | Proportion of Infectious being asymptomatic | 0.76 | - | King et al. (2008) |
| $e_1$ | Death rate due to asymptomatic | 0 | $\frac{1}{\text{year}}$ | King et al. (2008) |
| $e_2$ | Death rate due to symptomatic | 0.24 | $\frac{1}{\text{year}}$ | King et al. (2008) |
| $\gamma_1$ | Recovery Rate of asymptomatic | 0.15 | $\frac{1}{\text{day}}$ | Hendrix (1971) |
| $\gamma_2$ | Recovery Rate of symptomatic | 21.6 | $\frac{1}{\text{year}}$ | King et al. (2008) |
| $\eta_1$ | Shedding rate of asymptomatic | 0.5 | $\frac{1}{\text{day}}$ | Levine et al. (1988) |
| $\eta_2$ | Shedding rate of symptomatic | 50 | $\frac{1}{\text{day}}$ | Levine et al. (1988) |
| $\kappa_{H_i}$ | Half saturation constant of hyper-infectious | $10^6/700$ | $\frac{\text{cells}}{\text{ml}}$ | Hartley et al. (2006) |
| $\kappa_{L_i}$ | Half saturation constant of Less-infectious | $10^6$ | $\frac{\text{cells}}{\text{ml}}$ | Hartley et al. (2006) |
| $\beta_{H_i}$ | Ingestion rate of hyper-infectious | 0.15 | $\frac{1}{\text{week}}$ | Codeço (2001) |
| $\beta_{L_i}$ | Ingestion rate of less-infectious | 1.5 | $\frac{1}{\text{week}}$ | Codeço (2001) |
| $w$ | Immunity waning rate | 0.7 | $\frac{1}{\text{day}}$ | Longini et al. (2007) |
| $\delta$ | Bacteria death rate | 1/30 | $\frac{1}{\text{day}}$ | Hartley et al. (2006) |
| $\chi$ | Bacteria transition rate | 5 | $\frac{1}{\text{day}}$ | Hartley et al. (2006) |
| $a_1$ | Migration rate from city 1 to city 2 | varied | $\frac{1}{\text{day}}$ | - |
| $a_2$ | Migration rate from city 2 to city 1 | varied | $\frac{1}{\text{day}}$ | - |

The stability analysis indicates that a cholera outbreak can stay positive in one city until all the population gets into its recovery class, on the other hand, the disease stays zero in the other city. See Figures 2, 3, and 4 for the visualization of the human population in these cities. These figures are obtained using the parameter values given in Table 2 and different values of the migration rates *a*_1_ and *a*_2_. When I obtain the plot of human populations in Figures 2, 3, and 4, I considered three different scenarios for population sizes in cities of the metapopulation.

**Figure 2:**
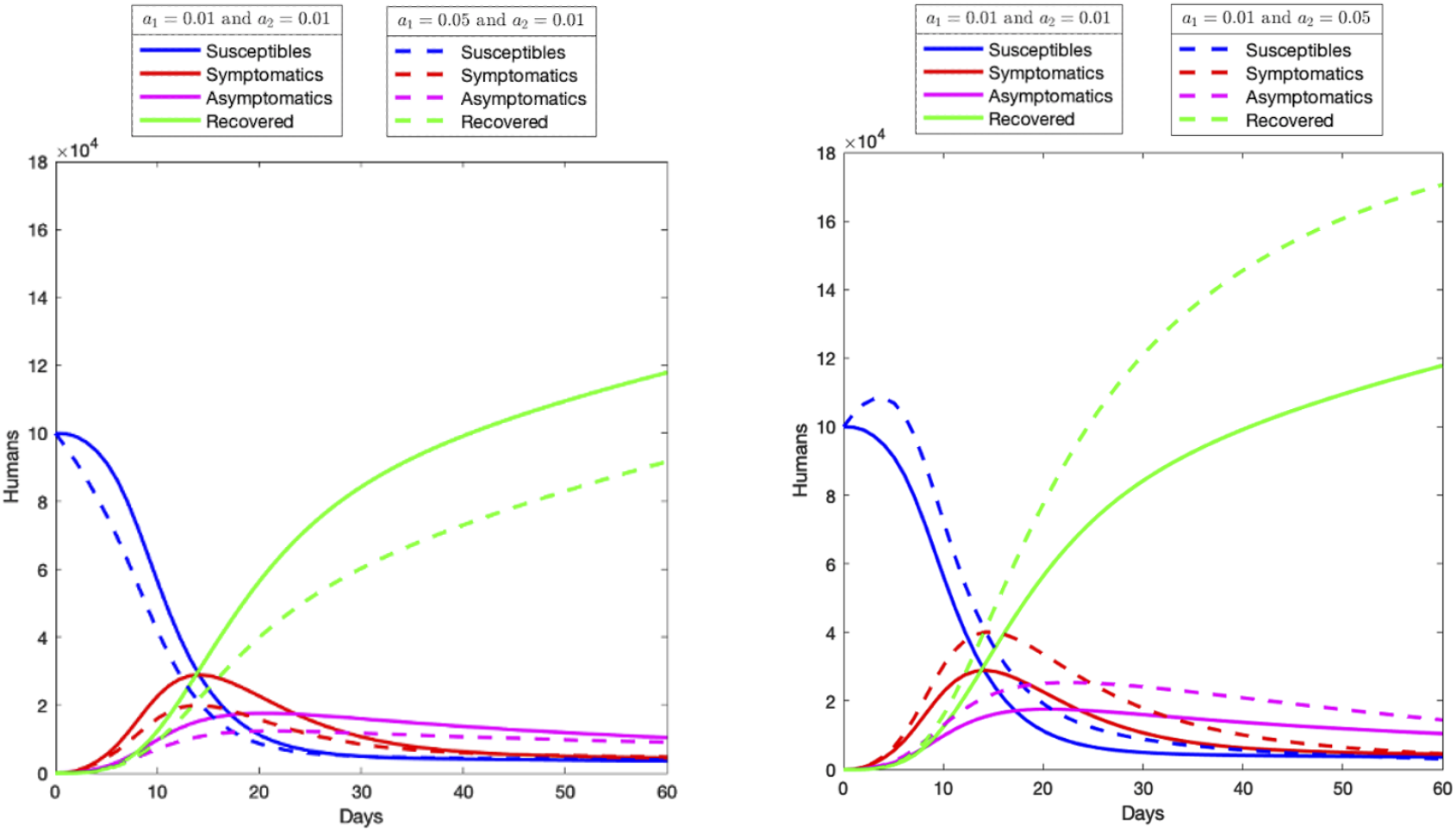
The number of susceptible, asymptomatic infected, symptomatic infected and recovered individuals in city 1 for the scenario 1.

**Figure 3:**
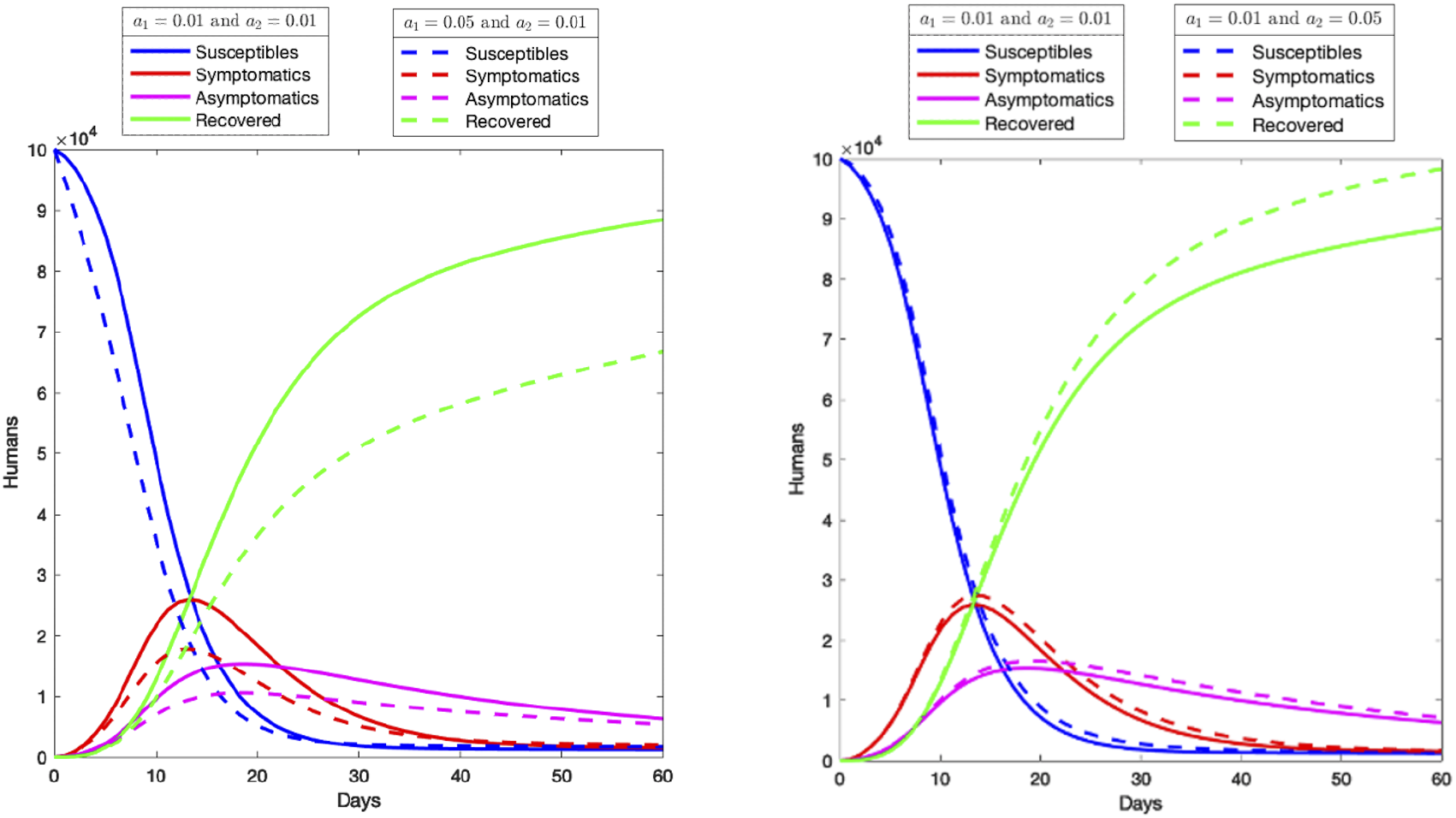
The number of susceptible, asymptomatic infected, symptomatic infected and recovered individuals in city 1 for the scenario 2.

**Figure 4:**
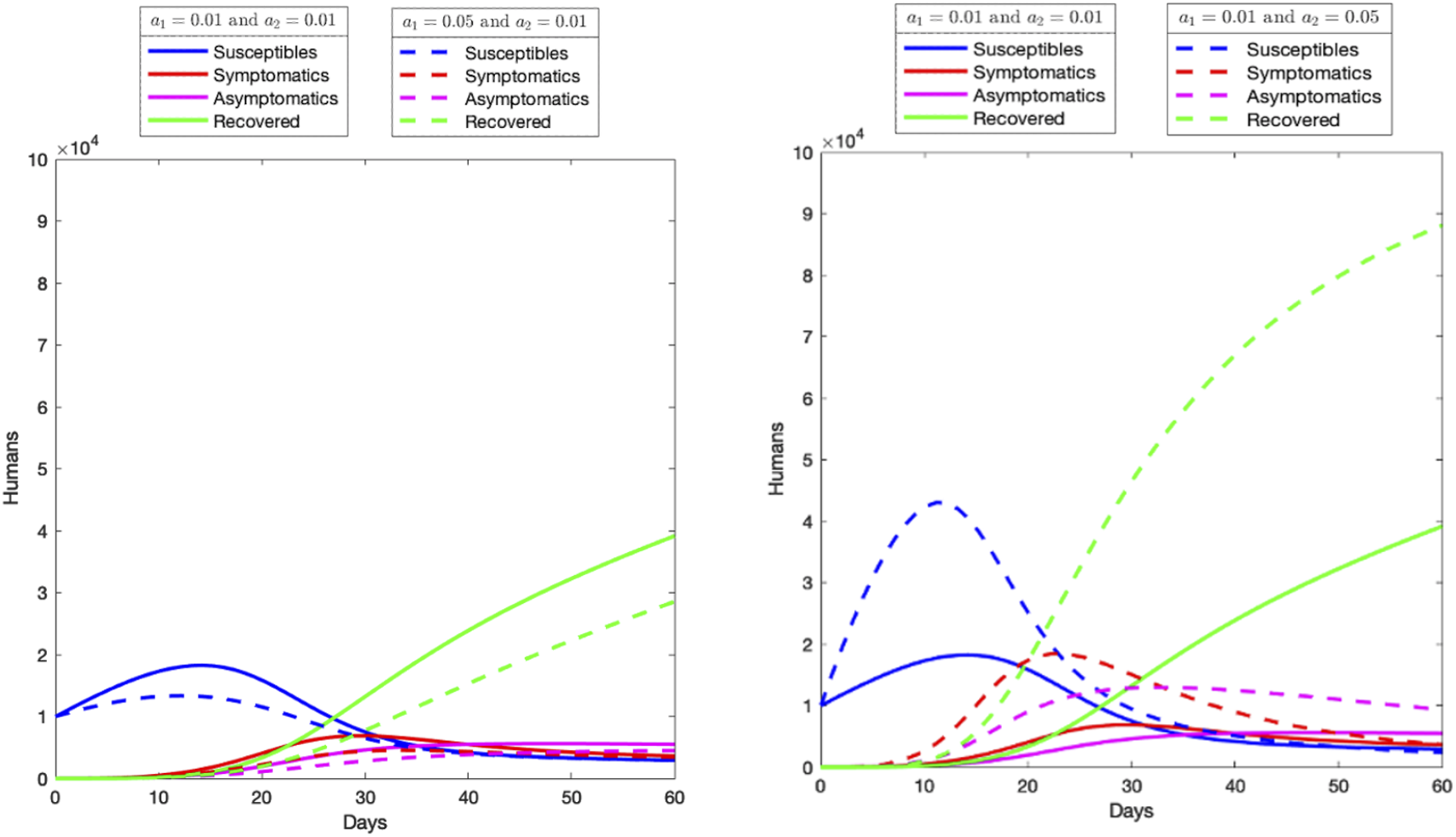
The number of susceptible, asymptomatic infected, symptomatic infected and recovered individuals in city 1 for the scenario 3.

**Scenario 1:** In this scenario, it is assumed that the human population in both cities has the same size of 100000 individuals. The effect of migration between these two cities is investigated when the migration rates *a*_1_ and *a*_2_ are varied from 0.01 to 0.05 (See Figure 2).

The left plot in Figure 2 shows that the number of asymptomatic and symptomatic infected cases decreases if more people move out of city 1 than move into city 1 during the 60 days. Also, we see that the number of asymptomatic and symptomatic infected cases increases when more people move to city 1 (See the right plot in Figure 2).

**Scenario 2:** In this scenario, it is assumed that the human population size is 100000 individuals for city 1 and 10000 individuals for city 2. The effect of migration between these two cities, especially the effect of migration in city 1 is examined by varying the migration rates *a*_1_ and *a*_2_. If we compare this scenario with the scenario 1, we see fewer increases in the number of asymptomatic and symptomatic infected cases in the right plot in Figure 3. It is mainly due to having 10 times less population in city 2 as compared to city 1 in this scenario.

**Scenario 3:** This scenario is vice versa of the scenario 2. Now, I assume that city 1 has 10000 individuals but city 2 has 100000 individuals. In this case, we see more increases in the number of asymptomatic and symptomatic infected cases in city 1 when we increase the migration rate *a*_2_ from 0.01 to 0.05 (See the right plot of Figure 4). it is mainly due to not only having more people moving into city 1 from city 2 but also having 10 times less population size in city 1 as compared to city 2.

These three scenarios are valid, especially for the cities where seasonal workers move from one city to the other city for a short time period (the period is assumed to be two months in this study). Such short-time movements may also occur due to some touristic purposes, natural disasters, or wars.

## 6 Conclusions

This study shows that the basic reproductive number, *R*_0_ obtained for two isolated cities also depends on population migration rates *a*_1_ and *a*_2_, and any increases or decreases in these rates directly affect the basic reproductive number. Thus, such migrations can not be ignored in the control of cholera outbreaks.

In addition, the existence of endemic equilibrium points is investigated. This investigation shows that when the endemic equilibrium points are stable, then the infected individuals can reach zero in one city, but the infected individuals still can exist in the other city for a cholera outbreak.

In the three scenarios given in section 4, the effect of migration rates *a*_1_ and *a*_2_ on asymptomatic and symptomatic infected cases are investigated (See Figures 2, 3, and 4). This investigation indicates that the number of asymptomatic and symptomatic infected cases can increase or decrease depending on the population sizes of cities and the migration rates *a*_1_ and *a*_2_. These increases (or decreases) change significantly depending on the difference in population sizes of cities.

## Data Availability

No data is used in this study

